# *“Feeding was such a huge element of how my illness manifested itself, it can’t just be me.”*: A qualitative study of infant feeding experiences and support in women with severe mental illness

**DOI:** 10.1101/2025.09.16.25335858

**Authors:** Natasha Baker, Claire A. Wilson, Debra Bick, Lousie M. Howard, Hannah Rayment Jones, Ioannis Bakolis, The infant feeding and mental health study working group, Tayana Soukup, Yan-Shing Chang

**Affiliations:** King’s College London; South London and Maudsley NHS Foundation Trust; University of Warwick; Imperial College London

**Keywords:** Perinatal, Women’s Mental Health, Mental Illness, Breastfeeding, Supplementary Feeding, Qualitative Research

## Abstract

**Background:** There are many benefits of breastfeeding to the physical and emotional wellbeing of mothers and babies but very little is currently known about the infant feeding experiences of women with severe mental illness (SMI). Due to a paucity of evidence in this area, women’s individual support needs are largely unknown.

**Aim:** To explore how women with SMI experience infant feeding and their support needs with this.

**Methods:** Semi structured interviews were conducted online and in person with 20 women.

Interviews were audio-recorded, transcribed and anonymised. Reflective thematic analysis was used to analyse the data.

**Findings:** Four key themes were identified; (1) The intersection between infant feeding and mental health; (2) Infant feeding support from maternity services; (3) Infant feeding preparation; and (4) Specific considerations for women with SMI. The concept of ‘joined up thinking to support infant feeding and mental health’ draws the themes together and is marked by lack of support specific to the individual needs of women with SMI.

**Discussion:** Findings revealed how SMI can manifest itself in beliefs and emotions related to infant feeding. Experiences of support highlighted the need for better anticipatory guidance around infant feeding for women with SMI, including contexts specific to perinatal SMI like psychiatric inpatient settings, psychotropic medication use, and the challenges associated with sleep deprivation.

**Conclusions:** Negative experiences with infant feeding and poor support have a significant impact on women with SMI. Support should focus on the emotional and practical demands of breastfeeding and how women can manage this alongside their illness.

## 1. Introduction

Severe mental illness (SMI) includes psychotic and mood disorders such as schizophrenia, bipolar disorder and severe depressive disorders. SMI often causes severe functional impairment and requires intensive input by mental health services^(1)^. The requirement for mental health services, rather than diagnosis was used to define SMI in this study. In a perinatal context, this includes women with a pre-existing illness who become pregnant, affecting around 2.8% of women universally, and those who first develop a severe psychiatric illness in pregnancy or postpartum, affecting 1-2 per 1000 births worldwide^(2)^.

SMI in the perinatal period is associated with poor birth outcomes and obstetric complications^(3)^ and women with SMI are less likely to initiate and continue breastfeeding^(4, 5)^. Breastfeeding is a public health priority, the benefits of which are well established^(6)^. UNICEF and The World Health Organisation (WHO) recommend exclusive breastfeeding for the first six months, and continued up to two years or longer alongside complementary foods^(7)^. However, high income countries fall short of these recommendations with 1in5 babies never receiving breastmilk^(7)^ and just 55% continuing to breastfeed at six weeks^(8)^. Studies exploring women’s experiences of breastfeeding suggest that although women are aware of the health benefits, they are often surprised by the challenges of breastfeeding and do not feel prepared to manage these challenges^(9)^.

Breastfeeding can be particularly challenging for women with SMI due to psychotropic medication needs and the importance of protecting sleep^(10)^. However, there is limited evidence exploring the IF experiences of women with SMI. Some previous studies have focused on women with opioid dependence^(11)^, those who are neurodivergent^(12)^ and the infant feeding (IF) perceptions of women with eating disorders (ED)^(13)^. There is more literature exploring how postnatal depression (PND) impacts women’s experiences of IF^(14)^ but evidence is limited in those with a clinical diagnosis of severe depression and other SMIs^(4)^.

Despite the benefits of breastfeeding and the potential to reduce health disparities for women with SMI and their infants, due to a paucity of evidence in this area women’s individual IF support needs remain unclear^(4)^. This study therefore aimed to explore the infant feeding (IF) experiences and support needs of women with SMI.

## 2. Methods

### 2.1. Design

A qualitative study using semi structured interviews was undertaken as part of a mixed methods study investigating IF in the context of SMI. The quantitative phase which will be reported separately was concurrent but independent to this qualitative study and compared infant feeding outcomes in women with and without SMI. Critical realist (CR) ontology and constructivist epistemology were adopted when collecting and analysing the data. CR recognises that knowledge is subject to historical, cultural, and social contexts^(15)^, whilst constructivism acknowledges an individual’s experiences and social contexts within the construct of truth^(16)^. The data collection and analysis methods detailed below support these philosophical views by acknowledging a reality independent from the researchers’ own perceptions. A reflexive journal was kept throughout data collection and reflective discussions within the research team helped to critically evaluate the researchers’ influence on the research, acknowledging individual assumptions and potential for bias. For example, NB is a midwife and mother with experience of breastfeeding and PND which we acknowledge may have influenced how participant interviews were perceived. Other members of the research team are also parents and come from multidisciplinary clinical and academic backgrounds and/or have clinical experience of caring for women with SMI in the perinatal period.

### 2.2 Setting

This study was carried out in the UK where pregnant and postpartum women access publicly funded national health services, combining primary care with maternity services. Women with SMI in the UK are usually seen by both maternity and secondary mental health services in the perinatal period, typically community perinatal mental health (PMH) services. Secondary services provide dedicated care by clinicians specialising in mental health, such as psychiatrists, psychologists and specialist nurses.

### 2.3 Participants and recruitment

Women were primarily recruited through purposive sampling at two PMH services, based in London and central England between 2022 and 2023. Women were also recruited through study posters displayed in clinical areas and by the charity, *Action Postpartum Psychosis.* Purposive sampling aimed to achieve maximum variation of mental health diagnosis and experiences of IF. Eligible women were initially approached by a health care professional from their perinatal team and later contacted by NB (See figure 1 for recruitment procedures). The inclusion criteria were women ≥ 18 years, who had a livebirth up to five years prior and received (any) secondary mental health care in the UK during pregnancy and/or the first three months postpartum. Up to five years was chosen to be as inclusive as possible of those wanting to share their experiences, whilst being mindful that recall could be influenced by time and infant development, though evidence suggests women remember their childbirth experiences accurately at 5 years postpartum^(17)^

**Figure 1:**
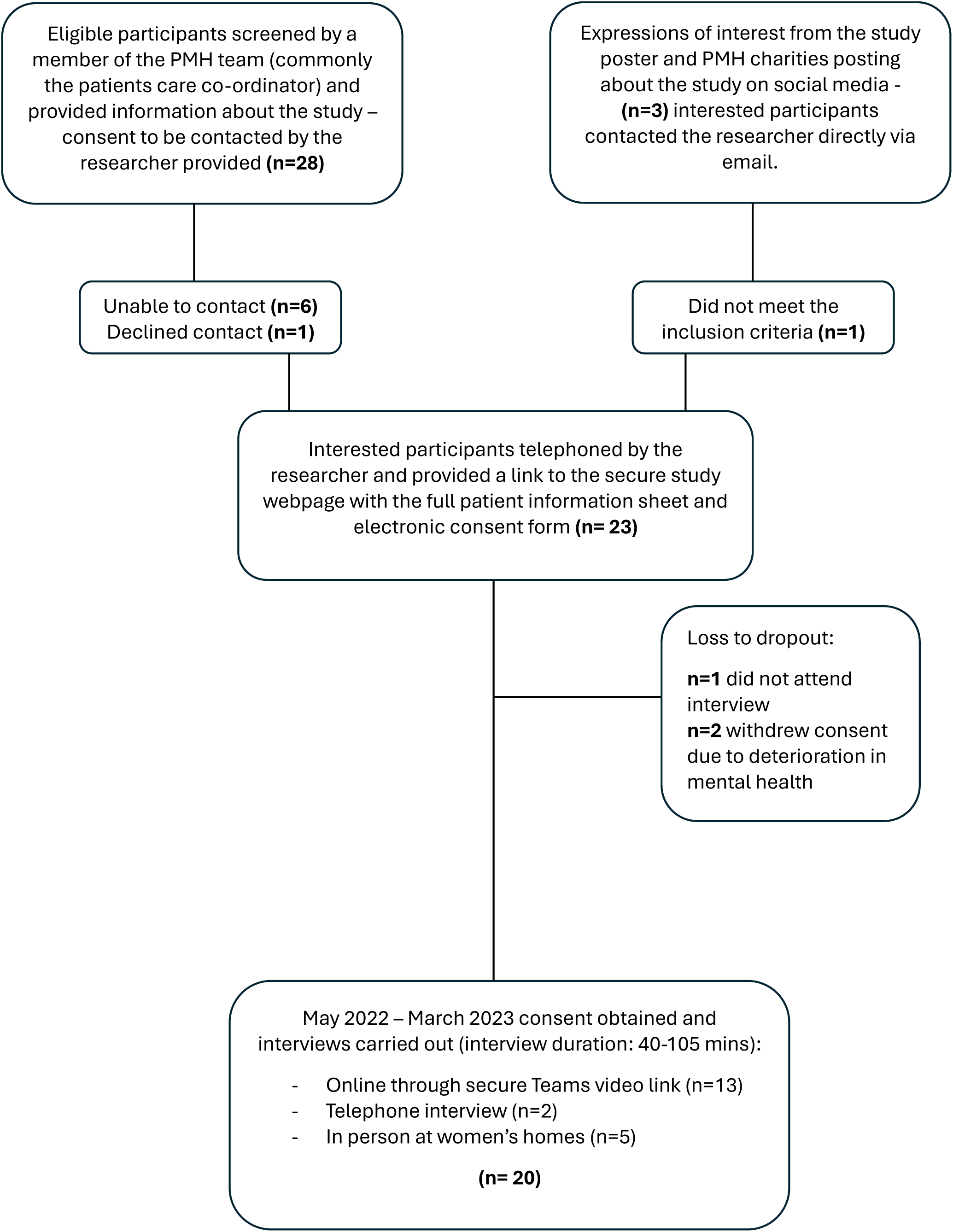
Recruitment flowchart.

### 2.4 Ethics

Ethical Approval was sought and granted in February 2022. All participants provided written informed consent prior to taking part in the study. All procedures contributing to this work comply with the ethical standards of the relevant national and institutional committees on human experimentation and with the Helsinki Declaration of 1975, as revised in 2008. Perinatal mental health and infant feeding can be emotionally triggering to discuss. Emotional wellbeing of participants was monitored throughout the interview. Participants were provided with written information signposting to relevant sources of emotional support after the interview and a distress protocol was in place if anyone required additional support.

### 2.5 Data collection

NB conducted all interviews either online through Microsoft Teams or in-person at women’s homes, dependent on participant preference. Interviews were recorded (through Teams and digitally), anonymised and transcribed verbatim by an independent transcription company. The interview guide was developed based on findings from existing literature^(10, 14, 18)^, clinical experience of the authors (NB, DB, CAW & LMH) and advice from the study’s patients and the public advisory (PPA) working groups. PPA groups include women with lived experience of perinatal SMI, carers of people with SMI and IF peer supporters. Two pilot interviews were also carried out with members of the PPA groups to test the topic guide. Participants’ characteristics (Tables 1) were collected using standardised forms. Data collection continued until there was adequate information power, defined as, the richness, variation and relevance of the data to the research objectives^(19)^.

**Table 1.**
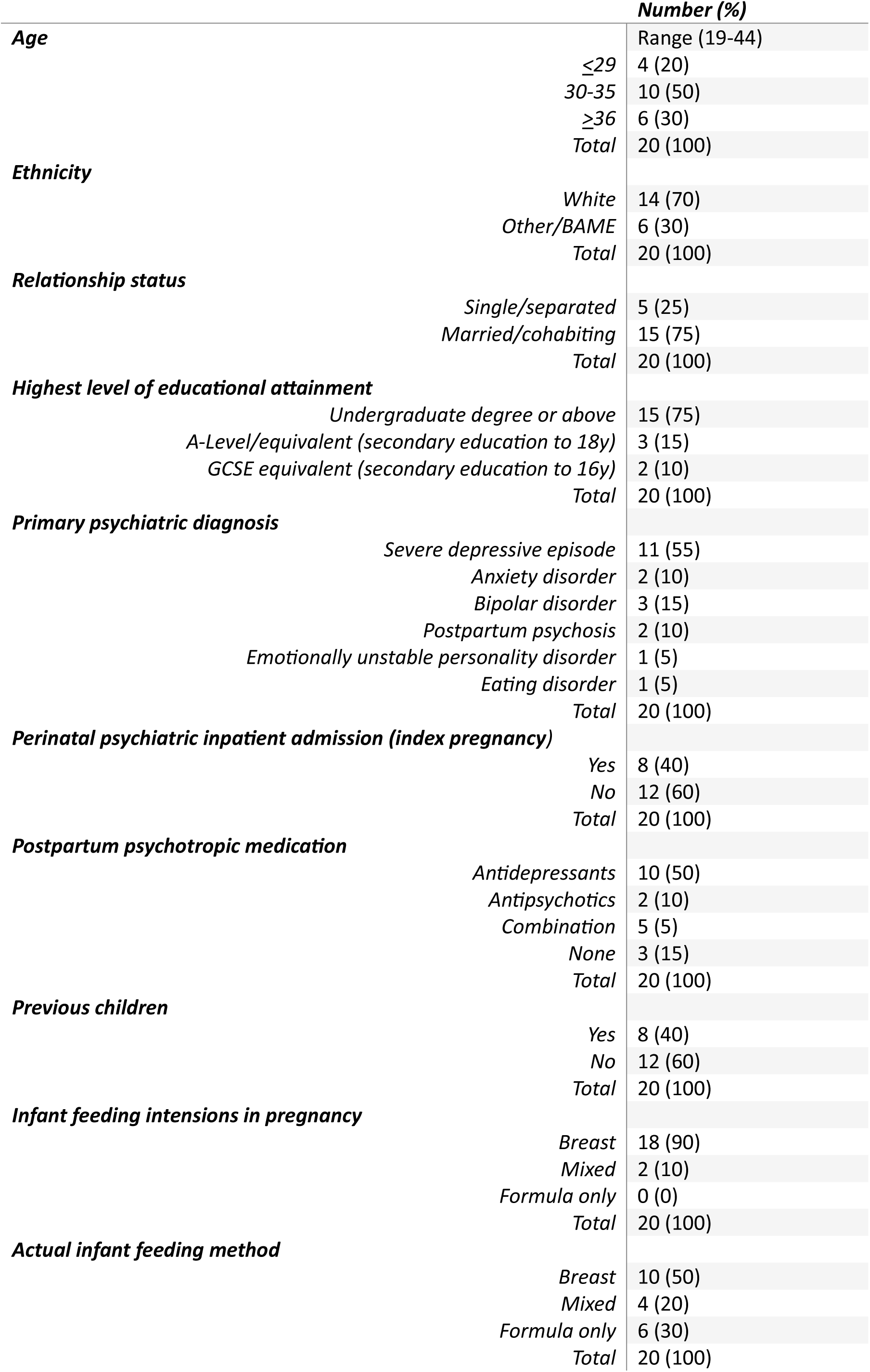
Participants Characteristics.

| **Table 1. Participants Characteristics** |  | **Number (%)** |
| --- | --- | --- |
| **Age** |  | Range (19-44) |
|  | ≤29 | 4 (20) |
|  | 30-35 | 10 (50) |
|  | ≥36 | 6 (30) |
|  | Total | 20 (100) |
| **Ethnicity** | White | 14 (70) |
|  | Other/BAME | 6 (30) |
|  | Total | 20 (100) |
| **Relationship status** | Single/separated | 5 (25) |
|  | Married/cohabiting | 15 (75) |
|  | Total | 20 (100) |
| **Highest level of educational attainment** | Undergraduate degree or above | 15 (75) |
|  | A-Level/equivalent (secondary education to 18y) | 3 (15) |
|  | GCSE equivalent (secondary education to 16y) | 2 (10) |
|  | Total | 20 (100) |
| **Primary psychiatric diagnosis** | Severe depressive episode | 11 (55) |
|  | Anxiety disorder | 2 (10) |
|  | Bipolar disorder | 3 (15) |
|  | Postpartum psychosis | 2 (10) |
|  | Emotionally unstable personality disorder | 1 (5) |
|  | Eating disorder | 1 (5) |
|  | Total | 20 (100) |
| **Perinatal psychiatric inpatient admission (index pregnancy)** | Yes | 8 (40) |
|  | No | 12 (60) |
|  | Total | 20 (100) |
| **Postpartum psychotropic medication** | Antidepressants | 10 (50) |
|  | Antipsychotics | 2 (10) |
|  | Combination | 5 (5) |
|  | None | 3 (15) |
|  | Total | 20 (100) |
| **Previous children** | Yes | 8 (40) |
|  | No | 12 (60) |
|  | Total | 20 (100) |
| **Infant feeding intentions in pregnancy** | Breast | 18 (90) |
|  | Mixed | 2 (10) |
|  | Formula only | 0 (0) |
|  | Total | 20 (100) |
| **Actual infant feeding method** | Breast | 10 (50) |
|  | Mixed | 4 (20) |
|  | Formula only | 6 (30) |
|  | Total | 20 (100) |

### 2.6 Data analysis

Data analysis was based on the six stages of Braun and Clarke’s reflexive thematic analysis (TA)^(16, 20),^ each phase of the analysis is described in more detail in table 2. Inductive TA was used, meaning themes were established through participant’s narratives, rather than a predefined criteria, which is appropriate for giving voice to marginalised groups^(16)^. Data were analysed using NVivo 14. A second independent coder (YSC analysed 10% of the transcripts, chosen at random and any disagreement in data interpretation was discussed. All findings were reviewed by and discussed with the studies PPA groups who were in agreement.

**Table 2.** Phases of reflexive thematic analysis based on Braun & Clarke (2006)

|  |  |  |
| --- | --- | --- |
| 1 | Familiarisation with the data | A reflexive diary was kept throughout the research process and during interviews to improve rigour, XX made continuous notes of her initial interpretation of women's narratives, and these were discussed with other authors. Data cleaning was carried out by listening to audio recordings whilst reading transcripts and making any amendments necessary, allowing familiarization and emersion in the data. |
| 2 | Generating initial codes | Interesting parts of the data which most applied to the research aims and objectives were coded by XX using NVivo software. Consensus coding was carried out by XXX who also coded a random 10% of the transcripts. Any disagreement was discussed between XX and XXX. If consensus could not be reached, this was discussed with the other authors. |
| 3 | Searching for themes | XX condensed codes into categories which became potential themes and sub-themes. |
| 4 | Reviewing themes | Coded extracts were reviewed against the themes, sub-themes and the entire dataset. Themes and sub-themes as well as coded extracts were sense checked with XXX. A theme map and summary of findings was then generated and presented to the studies PPA groups. |
| 5 | Defining and naming themes | XX further condensed and reorganised sub-themes under the four key, overarching themes and XXX sense-checked this. These were reviewed by all authors. |
| 6 | Producing the report | A lay summary of findings was produced and sent to participants who were invited to feedback their thoughts and validate the findings if they wished. No amendments were needed.<br>The main findings were written up by NB and reviewed by all authors. |

## 3. Findings

Most participants reported previous problems with their mental health (n=18), ranging from common mental disorders such as anxiety and depression, to chronic SMI. The predominant diagnosis was a mood disorder (n=14). Five women reported previous psychiatric inpatient admissions in their lifetime but for most, this was the first time they had required any type of secondary mental health care. Six women required admission to a psychiatric Mother and Baby Unit (MBU) and two were admitted to a general psychiatric ward (one subsequently transferred to an MBU). Several other women also required input from home treatment teams (HTT) or psychiatric liaison services in emergency departments. Most breastfed (n=14), though not exclusively.

Breastfeeding exclusivity and duration were not formally recorded to be sensitive to participants’ experiences by avoiding excessive focus on feeding method.

Four key themes were identified through thematic analysis and are visually presented in figure 2. The concept *‘joined up thinking to support IF and mental health*’ brings the themes together and is marked by a disconnect in services to support the IF support needs of women with SMI; *“It’s like either or, and there’s no one that really deals with both [infant feeding support & mental health]” (#12).* Additional quotes and their corresponding themes and sub-themes can be found in table.3.

**Figure 2:**
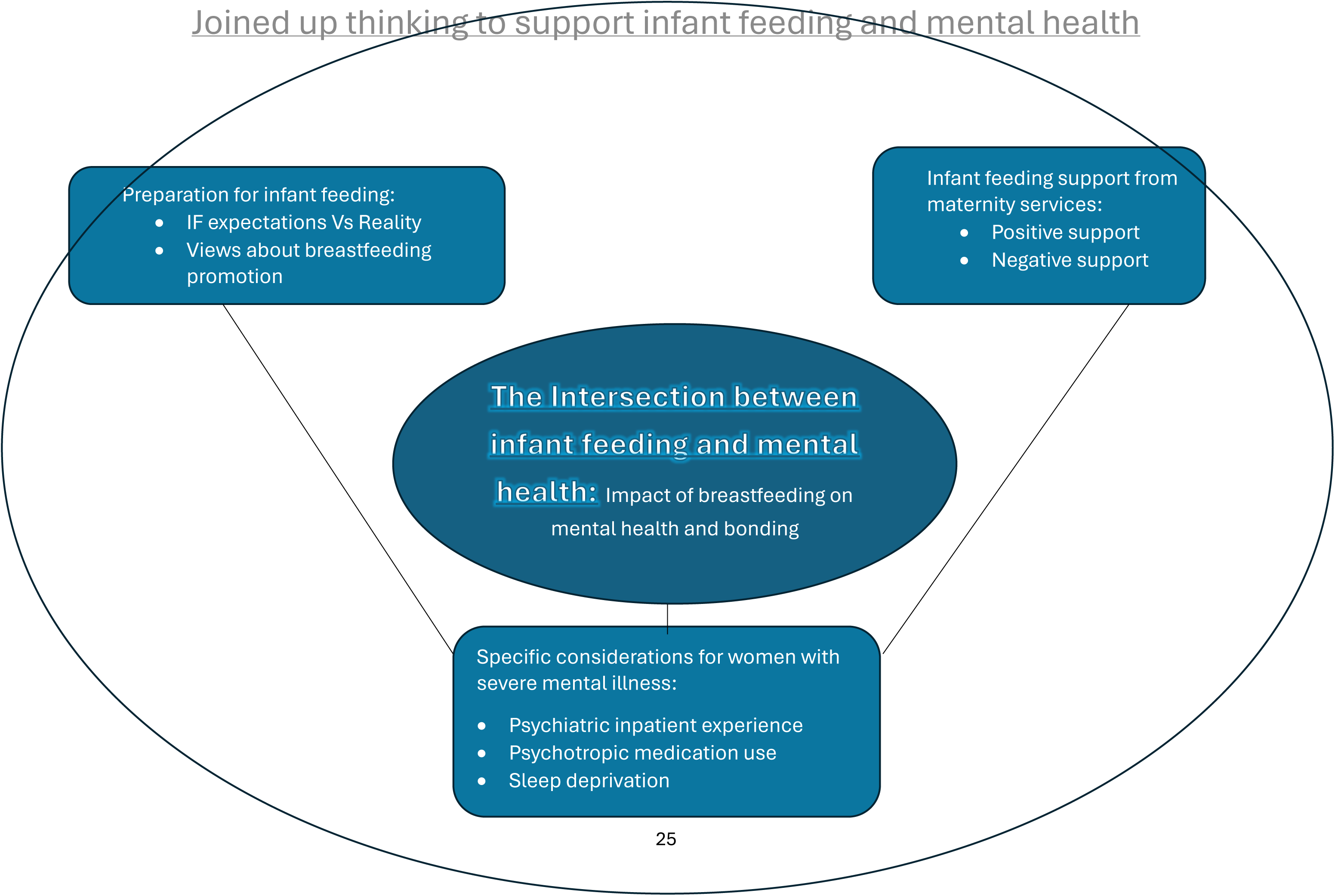
Theme map

### 3.1 Theme 1: The intersection between infant feeding and mental health

All participants described their emotions linked to IF and the experience of breastfeeding was described as intrinsically linked to their mental health:

> *“I think one of the ways my mental health problem manifested is through my absolute…I had to feed him, and it wasn’t for the normal reasons that people want to breastfeed, it was for bizarre, unhealthy mental health [reasons]” (#14)*

#### 3.1.1 Impact of breastfeeding on mental health and bonding

For some, breastfeeding was ‘*the only positive thing in a really bleak time’ (#16)* and protective of intrusive thoughts relating to suicide:

> *‘In times when I’ve desperately wanted to end my life breastfeeding has been like a small thread that has tied me to the earth’ (#19)*

The intimacy of breastfeeding was described as *‘grounding’,* helping women during episodes of extreme despair. Women with EDs also felt breastfeeding had helped them to minimise eating behaviours associated with their illness:

> *“Obviously there is the calories that’re used up that sort of help with the anxiety.” (#18)*

Several women found bonding with their baby challenging but when breastfeeding had gone well, the intimacy helped to overcome this and was a hugely motivating factor for breastfeeding. For several, breastfeeding was *‘the one thing that I could do that nobody else could do.’ (#18)* and helped reinforce the maternal-infant bond:

> *“…But then research had come in with the next baby that said, actually, no, this has been proven to be safe, you can breastfeed and I really enjoyed it, so I enjoyed that closeness. It really did for me personally intensify that bond.” (#20)*

However, for one participant, this had a detrimental impact on bonding as she felt feeding was her only role. This progressed to feelings of inadequacy as a mother and a subsequent suicide attempt.

Difficulties establishing breastfeeding compounded challenges with bonding:

> *“I was so much more focused on like my milk supply and not being able to do it than like any of the like bond with [baby].” (#3)*

Breastfeeding difficulties were associated with obsessive thoughts and behaviours: *“I just became like absolutely, I don’t know, obsessed with it working and with tracking it.” (#1).* Which was linked to compulsion to continue despite difficulties: *“My compulsion to keep feeding even though I was in huge amounts of pain, it definitely wasn’t normal and had I been in a better frame of mind I wouldn’t have struggled on with breastfeeding.” (#14)*. Those experiencing breastfeeding difficulties described negative thoughts and ruminations about breastfeeding, in particular failure, shame and guilt:

> *“All my NCT mums which just whip their boobs out and I felt like such a failure…Why can they do it and I can’t. They all can do it. I’m in the minority. I failed. I’m not good enough.” (#2)*

> *“…Embarrassed about making a bottle in public cos I thought everyone would be like judging me”’ (#3)*

### 3.2 Theme 2: Infant feeding support from maternity services

#### 3.2.1 Positive support

Family, social and peer support was central to women’s experience. Peer support was accessed formally through PMH teams and informally through new friendships with other mothers. Women valued shared experiences, *“Knowing that I’m not the only one who’s been in that position” (#13)* and particularly appreciated the opportunity to connect with other mothers living with mental illness:

> *“I communicate more, and hearing how they found it compared to how I found it and it was like, yeah, it really is different when you’ve got mental health or anxiety.” (#8)*

Similarly, women valued individualised support; “I think it’s just that feeling of having somebody that’s like, we are with you and supporting you and your feeding journey, whatever that may be.” (#11) and wanted permission to stop breastfeeding when it was impacting their mental health; “I wanted a health care professional to give me permission and say it’s okay, just stop. You’re not a failure if you stop.” (#2). One participant experienced difficulties breastfeeding due to sedating side effects of her medication. She received support from a midwife who facilitated her breastfeeding intentions whilst also supporting and understanding her mental health (Table 3).

**Table 3:** Additional quotes.

| Theme & sub-theme | Data example (participant quote): |
| --- | --- |
| **Theme 1: The intersection between breastfeeding and mental health** |  |
| Impact of breastfeeding on mental health & bonding | *"When I was like, very unwell, the only thing that was keeping me alive was the fact that he needed to feed from me, he wouldn't take a bottle" ... "I just remember thinking like as soon as he doesn't need me to feed him like, that's it, I'm not gonna be here anymore" (#14)*<br>*"I think it's been positive, and I think it's helped my, issues." (#10)*<br>*"I was like why are you hurting me? You shouldn't, it shouldn't. It wasn't a positive connection that I had with him." (#14)* |
| **Theme 2: Infant feeding support from maternity services** |  |
| Positive support: Peer support | *"...A really supportive women's group, like a mum and baby group. But we don't do baby singing, we talk about our favourite books, depression, breastfeeding trauma, sleep." (#6)* |
| Individualised support | *"She knew I wanted to breastfeed. She was training herself to be a mental health midwife. She came into my section with me, and she was phenomenal. And she just sat with me when I was drowsy and everything, she just sat with her hands on my shoulder, you know, come on, shall I take the baby now, for you? And she'd take the baby and put her in the cot for me. She'd come in and check now and then... could there be a resource where there is a little team that does that, that helps women who are on psychotic medication?" (#20)* |
| Negative support: contradictory advice | *"Health visitors, midwives, doctors, nurses on the ward, literally everyone has their own opinion. It was a mess." (#17)* |
| Difficulties getting support with IF | *"I couldn't take my medication in hospital in the end, because I couldn't safely bottle feed him either. So I had to not take my medication." (#20)*<br>*'Like no one helps you with mastitis.' (#2)* |
| Unheard | *"I didn't feel listened to, and I didn't feel like she was actually very interested in my specific set of issues" (#17)* |
| **Theme 3: Preparation for breastfeeding** |  |
| IF expectations Vs reality | *"I didn't think it was going to be that much more challenging because I thought, oh, I know exactly what to expect, you know, done this before but it was different after birth, I must say." (#15)*<br>*"It would have been useful to know how difficult it was gonna be" (#14)* |
| **Theme 4. Specific considerations for women with SMI** |  |
| Psychiatric inpatient experience and IF | *"I'd say it was like I hadn't given birth until I got to the MBU and then it was all about having given birth. It was like chalk and cheese" (#11)* |
| Psychotropic medication use and IF | *"I was like, is it okay for me to breastfeed her or not? Because I can't come off these medications, I won't be able to look after her at all. So it was a bit like, can somebody just make their mind up if I'm poisoning my baby or not." (#17)*<br>*"I think I thought that if it wasn't for, you know, 40 weeks of my medicine they wouldn't be screening you for all these things because I know you're fine and this is what I've caused that I have to stop putting this into your system." (#4)* |
| Sleep deprivation | *"It was months off before I was gonna get sleep, so kind of the emphasis on how important it was to get up. You get up at this time, you get up at this time, felt really quite actually kind of damaging" (#11)* |

Individualised support was more accessible for women who had continuity of midwifery carer. This was extremely valued by women in the study and trust was a key part of this. Women also appreciated not having to repeat their story and felt continuity helped minimise contradictory advice:

> *“But I think having everyone tell you something different because you’re seeing different people so I don’t know if it would help more, you actually have one person because then you’re not hearing other people’s opinions.” (#3)*

#### 3.2.2 Negative support

Even when there were positive aspects of care, participants felt let down by IF support, with some paying for private lactation consultants after leaving hospital. Key aspects of this were staff shortages and lack of time, insensitive, contradictory advice and the impact of the COVID-19 pandemic. Poor support compounded IF difficulties and mental illness:

> *“I think that emphasis (on breastfeeding), but without lack of support really was a major factor in the episode I had after I gave birth” (#1)*

> *“There was two midwives covering that whole ward, so that was how short of staff they were and because of that, obviously, breastfeeding is not a priority because keeping people alive and safe is so there was no support whatsoever.” (#20)*

One participant with bipolar disorder reported stopping antipsychotic medication as the side effects meant she could not properly care for her baby unsupported on the postnatal ward. Support was even less accessible later in the postpartum period and lack of compassion with mastitis was described:

> *“Just so little compassion for what an active like self-mutilation that is when you’re in that much pain to carry on feeding, and how that might change how you felt about your baby” (#19)*

Overall, women felt unheard and that their needs were secondary to the baby’s feeding needs.

> *“It was all very tailored around feeding him and getting him to put on weight and getting him feeding. There was never any talk to me about how I was coping with it all.” (#12)*

Women whose babies were not gaining weight were given ‘feeding plans’ which left no time for sleep or self-care; *“I wasn’t sleeping because I was too busy feeding and expressing, you know and it was literally the worst idea ever.” (#12)*. For one woman with twins and one twin very unwell in a children’s hospital, this ‘*regime*’ was unsustainable, and she went onto develop a severe postpartum psychosis; *“I remember coming in in kind of tears and I was just like I just can’t keep doing this. I think you know it was silly given our circumstances to have been following this regime.” (#11)*.

### 3.3 Theme 3. Infant feeding preparation

#### 3.3.1 IF expectations Vs reality

All participants planned to breastfeed in some capacity and motivations to breastfeed related to bonding and it ‘*being best’* for the baby. IF expectations were based on previous IF experiences, cultural expectations, including family’s and friends’ opinions, and plans for returning to work. However, the reality of IF was significantly different to their expectations:

> *“But then when I did start to breastfeed, I wasn’t prepared for the actual struggles I went through” (#7)*

Women wanted realistic guidance about breastfeeding including the emotional and practical challenges:

> *“I think there’s no focus on the emotional side of it. Like you are the thing keeping your baby alive, and no one else can do that if you are breastfeeding” (#3)*

A participant with older children had excellent insight into her own mental health and spoke of having ‘*realistic expectations of IF’,* planning to mix feed so her partner could support her at night:

> *“As a bipolar mother, I know that one of my biggest triggers is sleep deprivation and so I’m very lucky that I have [baby’s] dad, who can do night feeds” (#20)*

Others did not feel as prepared and wanted more advice specific to their mental health needs:

> *“I think the discussion should come earlier on in pregnancy with mothers who do have a history of mental health…have you thought about how this may impact you or how your mental health may be impacted by whether you choose formula or breastfeeding, those never came into conversation” (#4)*

#### 3.3.2 Views about breastfeeding promotion

Participants described breastfeeding promotion as unhelpful and inappropriate. They felt it implied risks of formula feeding and therefore felt under pressure to breastfeed:

> *“It shouldn’t be in a paediatric A&E, you know…Every time I saw one of those fucking posters that said breastfeeding halves the risk of SIDS, all my mind could see was bottle feeding doubles the risk of SIDS” (#6)*

Women described breastfeeding promotion as ‘*obsession*’ with breastfeeding but felt this did not correspond with their subsequent experience of breastfeeding support and that IF support was limited to only those who were breastfeeding:

> *“I think like there is far too much emphasis…it’s like this obsession with it but at the same time it’s just like go off and sort yourself out.” (#1)*

> *“I don’t think a lot of people would not breastfeed due to lack of not feeling like they know that breast milk is really good for their baby. I feel like the literature kind of makes that kind of assumption rather than you know, that then kind of where is the support?” (#11)*

> *“Pretty much the only conversations relating to feeding and food were about breastfeeding” (#13).*

### 3.4 Theme 4. Specific considerations for women with SMI

#### 3.4.1 Psychiatric inpatient experience and IF

One participant was first admitted to a general psychiatric ward, then transferred to an MBU described the experiences ‘*like chalk and cheese’*, receiving no breastfeeding support or help with baby visits on the general ward:

> *“I remember a woman taking me for a shower because my T-shirts kept getting soaked through because, you know, I was by the end expressing for twins”…“so I sometimes think to have gone from expressing for twins and expressing quite full, a lot of volume to then nothing that that could the hormone effect could potentially contributed to illness” (#11)*

> *“I’ve read in my notes that grandparents want to bring in [twin1], we’ve told them no.” (#11)*

MBU experiences were more positive; focus was given to mother-infant bonding as unlike general psychiatric wards, MBUs in the UK allow mother and infant to stay together on the ward; *“I just felt like I’m finally safe” (#11).* However, participants reported similar difficulties being separated from their baby at night (in a separate room of the MBU) and lack of understanding about how this could impact breastfeeding:

> *“So that suddenly meant that, you know, 10 hours of the day I wasn’t breastfeeding and I arrived and was very nervous about my supply and so asked for like a pump, which they did provide. But like it took a while, no one really knew how to use it.” (#1)*

Commonly nursery nurses cared for babies at night to protect maternal sleep, but women described still waking to check their babies:

> *’I’d be given hypnotics that, you know, were really strong, it knocked me out, but in spite of the effects of the medication, my mind would not rest until I’d wake up at the time that I’d be normally giving him the feed and I’d go and check up on him” (#16)*

Overall, women felt breastfeeding was deprioritised and poorly understood within the MBU:

> *“I could tell the mother and baby unit had been physically built and designed, and their processes designed around bottle fed babies and they just couldn’t really, consistently, you know in any meaningful way get their head around a demand led, breastfed baby” (#6)*

> *“In the kitchen is this big whiteboard and it says like, how all the babies are fed, so that all of the staff can help feed the babies and then like there were two children that were breastfed and like it just has nothing next to their names.” (#19)*

#### 3.4.2 Psychotropic medication use and IF

Although two participants reported conflicting advice about taking medication whist breastfeeding;

*“So it was a bit like, can somebody just make their mind up if I’m poisoning my baby or not.” (#17); “I felt like any midwife I did speak to felt unsure of their knowledge” (#4)*, others had good experiences seeking guidance from professionals, particularly perinatal psychiatrists:

> *“I really struggled with taking medication. So, [perinatal psychiatrist] was absolutely brilliant, she printed off everything for me, all of the research about like all of those different outcomes, and we went through like the lit reviews” (#19)*

Participant had concerns about the potential risks of taking medication to their baby and lack of evidence base, leading one participant to stop her medication in the postpartum:

> *“You know, there isn’t enough evidence and so I decided to come off my medication”(#4)*

#### 3.4.3 Sleep deprivation

Sleep deprivation was a focal aspect of women’s experiences; “S*leep and feeling overwhelmed was quite pivotal in that kind of crisis” (#1)* and was described as intrinsically linked with IF and mental health:

> *“I think there is an absolute gap that’s got to be recognised - that the feeding can absolutely contribute to a lack of sleep and lack of sleep can contribute to lots of mental health problems.” (#11).*

Some women experienced insomnia in the postpartum, struggling with ruminations about feeding in the middle of the night:

> *“I would just be up awake not being able to do anything and the mind just going literally like, when’s the next bottle” (#3)*

Some women found co-sleeping beneficial to protect their own sleep; *“We would co-sleep and that’s how I could then feed him, and then still maintain my sleep.” (#16)* but health professionals lacked understanding around this. Overall, maternity providers lacked understanding that sleep can impact mental health and mental health providers lacked insight into the perinatal context of sleep, particularly for breastfeeding mothers who were told *‘Just stop feeding’ (#12):*

> *“The biggest failing for new parents in my opinion is that there is just virtually zero support around infant sleep and more importantly, maternal and parental sleep.” (#6)*

> *“When they’re [HTT] talking about sleep and stuff, like getting sleep, like there’s no consideration for the fact that you’ve got a newborn at home” (#12)*

## 4. Discussion

Our findings highlight a fundamental link between IF and mental health. When breastfeeding went well, it was hugely positive and helped women through a very difficult time in their life, but breastfeeding difficulties were linked to obsessive thoughts and ruminations focused on failure and shame. Women highlighted a lack of IF support, particularly in the context of their mental health needs; women felt their needs were inferior to the baby’s and fundamentally unheard. Participants described a lack of adequate preparation to manage the emotional and physical demands associated with breastfeeding and wanted practical support with sleep deprivation.

Whilst prior research demonstrates an association between breastfeeding difficulties and subsequent mental health problems, particularly PND^(14, 21–24)^, ours is the first study to provide in-depth evidence of the intrinsic link between IF and SMI. For example, several women in our study had experienced suicidal ideations but had found the intimacy of breastfeeding grounding in these moments of despair and some felt the responsibility of breastfeeding stopped them acting on these thoughts.

This has not previously been demonstrated in the literature and was a key finding from theme one.

Women experiencing breastfeeding difficulties perceived their feelings about breastfeeding as obsessive, with associated compulsions to persist, and this being a catalyst to their mental illness. In an interpretive phenomenological analysis of women (without SMI) continuing to breastfeed despite difficulties, women positively re-framed their need to continue as ‘*determination and persistence’*^(25)^, demonstrating key differences in how women with and without mental illness perceive IF challenges.

Feelings of failure, guilt and shame if breastfeeding had been unsuccessful have been highlighted in other qualitive research, both in women with and without mental illness^(24, 26, 27)^. These studies highlight perceived and societal pressure to breastfeed and limited guidance on formula feeding which links to findings from our second theme. Poor IF support and perceived pressure to breastfeed have been shown to increase feelings of guilt and shame and are associated with anxiety and depression^(28)^. Our findings also align with a qualitative study of women with bipolar disorder who reported receiving inadequate formula feeding advice and feeling stigmatised for formula feeding^(29)^.

The evidence relating to theme two is concerning when considering what additional impact IF challenges and poor support may have on women with SMI.

Positive support was underpinned by individualised care, continuity of midwifery carer and peer support. Peer support for women with perinatal mental illness has previously been shown to be effective; stigma and isolation is counteracted by shared experiences and connection to peers is ‘*considered a valuable aspect of the journey towards recovery*’^(30)^. The benefits of IF peer support are well acknowledged (among women without SMI) and the World Health Organisation advocates it’s importance, with many areas of the UK now providing an IF peer support service^(31)^. Future research could explore the benefits of IF peer support to the experiences and IF outcomes of women with SMI.

Evidence from theme three demonstrated women in this study wanted realistic guidance around IF, based on the emotional and practical challenges of breastfeeding, and unbiased advice about formula and mix feeding. A recently published study investigating the impact of breastfeeding difficulties on maternal mental health also addresses the need for improved parental education about the realities of breastfeeding^(24)^. Addressing the balance between setting realistic expectations and sharing the many benefits of breastfeeding is complex, particularly for women with SMI who face unique IF challenges. However, our study and previous research in women without SMI indicate IF advice and support is bias towards breastfeeding and calls for a more balanced and family-centred approach to IF policy and support^(32)^. It is important that women with SMI are fully informed of the risks and benefits of breastfeeding, the potential challenges involved and offered alternative options, such as mix feeding. The preconception period presents an opportunity to provide valuable public health information, such as IF guidance, potentially improving outcomes for women with SMI and their infants^(33)^. Future research could explore the role of anticipatory IF guidance as a routine part of mental health preconception counselling.

Within theme four, women in our study felt psychiatric MBUs were hugely therapeutic and beneficial to themselves and their infants but were not set up to support breastfeeding. Previous research has shown that whilst women feel ‘*in good hands’* in MBUs, they require greater support with IF^(34)^. This includes, help to continue breastfeeding alongside psychiatric treatment such as protected sleep and psychotropic medication^(10)^. Our findings relating to psychotropic medication and breastfeeding were more positive than previous evidence^(4)^, as most women were happy with information provided. One explanation could be that women in our sample all had access to specialist advice from perinatal psychiatrists. However, women still experienced conflicting pressure about breastfeeding and the potential effects on their baby of taking medication. IF decisions for women with SMI require careful consideration of medication needs but should support breastfeeding in any capacity, dependant on maternal choice.

An important perception of women in our study related to the link between IF, sleep and mental health. Sleep disturbance is very common in both mental illness and in the postpartum period^(35)^.

Overall, women felt there was a lack of compassionate support for parental sleep and limited awareness about the impact this can have on mental health. Better support with postpartum sleep is required and professionals should offer solutions rather than minimise the impact of sleep deprivation^(36)^. Examples from our study include, sharing night feeds to aid a sufficient *‘block of sleep’* and providing advice around safe co-sleeping.

The present study reveals the complex IF support needs of women with SMI. We highlight some similarities and unique differences compared to literature in women without SMI. Support, including in the preconception period should focus on preparation for the emotional and practical challenges of breastfeeding, unbiased guidance relating to all types of IF and compassionate support, with women’s mental health needs at the centre. Future research and IF policy should include the use of psychotropic medication when breastfeeding, solution-based support with postpartum sleep and importantly, improvements to joined up working between maternity and mental health services. At its core, this involves mutual understanding of the importance of breastfeeding to women with SMI and appreciation of women’s mental health within this. Enhanced training is required for both maternity and mental health professionals to help them respond to the IF support needs of women with SMI, particularly in inpatient psychiatric settings.

### 4.1 Strengths and limitations

To our knowledge this is the first qualitative interview study to specifically explore the IF experiences of women with SMI. The study included a diverse sample recruited from across the UK, providing insights into different health service contexts, albeit the sample was mostly White, well-educated and married. The inclusion of several PPA members throughout the research process has ensured the research is relevant and meaningful to those with lived experience. A potential limitation of the study was that recruitment was mostly through health care providers, inviting a degree of selection bias, whereby providers select participants who they believe are more suitable for the research.

Despite this, information power was achieved within the sample as evidenced by the richness and variation of the data.

## 5. Conclusion

The IF experiences of women in our study highlighted that breastfeeding can be beneficial to the emotional wellbeing of women with SMI. However, women did not feel prepared for the emotional and physical challenges of breastfeeding and experiences of psychiatric inpatient environments revealed a lack of understanding about breastfeeding. Overall, women described a disconnect in services to adequately support IF and mental health, highlighting the importance of raising awareness regarding the complexities of IF for women with SMI.

## Consent for publication

Written informed consent was obtained from all participants in the study.

## Data availability

The datasets generated and analysed during the current study are not publicly available due to the privacy of the participants in the study and the sensitive nature of the data. Further inquiries can be directed to the corresponding author.

